# Female gender and knowing a person positive for COVID-19 significantly increases fear levels in the Cuban population

**DOI:** 10.1101/2021.03.14.21253561

**Authors:** Yunier Broche-Pérez, Zoylen Fernández-Fleites, Evelyn Fernández-Castillo, Elizabeth Jiménez-Puig, Dunia Ferrer-Lozano, Annia Vizcaíno-Escobar, Lesnay Martínez-Rodríguez, Reinier Martín-González, Boris C. Rodríguez-Martín

## Abstract

The objective of this study was to explore the relationship between sociodemographic factors and fear of COVID-19 in a Cuban population. A web-based study with a cross-sectional design was conducted. The sample comprised 1145 participants. To explore fear, the Fear of COVID-19 Scale was used. Our results suggest that women were more likely to experience medium to high levels fear compared to men. Additionally, knowing a person positive to COVID-19 significantly increases fear levels in Cuban participants.

## 1. Introduction

The fast spread of the SARS-CoV-2, and the measures to contain it, has lead to mental health problems around the world. Among the most frequent mental health problems related to COVID-19 reported to date are anxiety, insomnia, stress, depression, anger and fear (Rossi et al., 2020; Wang et al., 2020).

In Cuba to date (27/01/21), 23,439 cases positive for COVID-19 have been reported and 204 people have died because of this disease (Ministry of Public Health, 2021). However, there have been very few empirical studies evaluating the impact of the pandemic on the psychological well-being among Cubans. The three empirical studies published so far have evaluated the reactions of fear, anxiety, depression, stress and suicidal ideation (Arias-Molina et al., 2020; Broche-Pérez et al., 2020; Broche-Pérez et al., 2020). The results suggest a predominance of fear, anxiety and depression reactions in the Cuban population as the predominant psychological response to the pandemic. However, the conclusions of these investigations are based on relatively small samples ranging between 287 and 772 participants, making it difficult to obtain a more comprehensive perspective of the mental health of Cubans during the pandemic. In addition, studies that have included predictors have limited themselves to exploring the effect of gender (Broche-Pérez et al., 2020) and, for example, have not considered interpersonal proximity variables. For example, personal closeness with people positive for the disease, or at risk of becoming ill, could explain the differences in fear reactions to COVID-19. In this sense, the objective of this study is to explore the relationship between sociodemographic factors and fear of COVID-19 in a Cuban population.

## 2. Methods

### 2.1. Participants and Study Design

The sample of the study was made up of 1145 Cuban participants over 18 years of age. A web-based study with a cross-sectional design was conducted. The research was conducted between April and October 2020. The survey was developed using the free software Google Forms®. The study design was approved by the ethics committee of the Department of Psychology of the Universidad Central “Marta Abreu” de Las Villas, Cuba. The investigation was conducted in accordance with the Declaration of Helsinki of 1975, as revised in 2000. All participants provided informed consent prior to participating.

### 2.2. Measures

#### 2.2.1. Demographic Information

The survey collected demographic data such as gender and age (in years). Participants also answered the following questions: a member of family or friend has been confirmed with the disease? (“yes” or “no”); knew an infected person? (“yes” or “no”); residence in a place with confirmed cases of COVID-19? (“yes”, “no” or “I don’t know”) and country region (“west region”, “central region” or “east region”).

#### 2.2.2. The Cuban Fear of COVID-19 Scale (FCV-19S)

The Cuban FCV-19S (Broche-Pérez et al., 2020) assesses fear towards COVID-19 and was adapted from the scale developed by Ahorsu et al. (2020). The FCV-19S is a seven items scale with a five-item Likert-point response from 1 (*strongly disagree*) to 5 (*strongly agree*). For the Cuban population the Cronbach alpha coefficient was 0.80. The FCV-19S total scores range from 7 to 35. Higher scores indicate greater fear of covid-19.

### 2.3. Statistical analysis

The data were processed using SPSS/Windows, version 21. Descriptive statistics was used to explore participants’ characteristics. An independent-samples t-test was computed to compare fear reactions between participants, based on gender (male/female), member of family (or friend) positive for COVID-19 (yes/no) and knew an infected person (yes/no).

The Cohen’s *d* were calculated to estimate effect sizes in all comparisons (0.2, 0.5, and 0.8 were considered as small, medium, and large effect size, respectively (Cohen, 1988)). Demographic and social factors independently associated with levels of fear were investigated using multinomial logistic regression.

### 2.4. Data Sharing Statement

The current article includes the complete raw data-set collected in the study including the participants’ data set, syntax file and log files for analysis. All of the data files will be automatically uploaded to the Figshare repository.

## 3. Results

### 3.1. Characteristics of the sample

The age of the participants ranged between 18 and 81 years (*M*=30, *SD*±10.2). The age group with the most participants was the one between 18 and 30 years (59.7%) and women (68.5%) predominated. Most of the participants resided in places with positive COVID-19 cases (66.2%), however only 9.5% of those who completed the survey have had friends or family members infected. Of the total participants, 28.6% know someone (other than friends and family) who has been ill with COVID-19 and the participants from the central region of the country (75.1%) predominated. Medium (47%) and high (27.6%) levels of fear predominated in the sample.

### 3.2. Association between fear of COVID-19 and sociodemographic and exposure variables

In bivariate analyses, there was a significant difference in FCV-19S scores between males (*M*=17.11, *SD*=7.13) and females [*M*=20.42, *SD*=6.56; *t*(1143)=7.728, *p*<.001, *d*=.49]. Further, participants who endorsed “knowing someone infected by COVID-19” scored significantly higher (*M*=20.37, *SD*=6.62) compared to those who did not (*M*=18.98, *SD*=6.95; *t*(1143)=3.066, *p*=.002, *d*=.20). No other bivariate analysis was statistically significant.

### 3.3. Independent predictors of fear of COVID-19

*Female gender* significantly predicted whether they had middle or low fear to COVID-19, [*b* = −.98, Wald X^2^(1) = 39.09, *p* < 0.001] (table 1). The gender also significantly predicted whether people had high or low fear [*b* = −1.51, Wald X^2^(1) = 62.89, *p* < .001]. *Knowing an infected person* predicted whether they had middle or low fear to Coronavirus [*b* = −.90, Wald X^2^(1) = 19.92, *p* < 0.001] and also predicted high or low fear level [*b* = −.98, Wald X^2^(1) = 19.10, \**p* < 0.001].

**Table 1.** Demographic and social factors independently associated with fear of COVID-19

|  | B | SE | OR | CI 95%<br>Inf. | Sup. |
| --- | --- | --- | --- | --- | --- |
| **Middle vs. Low Fear<sup>a</sup>** |  |  |  |  |  |
| Intercept | .94 | .54 |  |  |  |
| **Age** |  |  |  |  |  |
| 18-30 years | -.41 | .29 | .66 | .38 | 1.17 |
| 31-45 years | -.25 | .30 | .78 | .43 | 1.41 |
| >45 years |  |  |  |  |  |
| **Gender** |  |  |  |  |  |
| Male | -.98 | .16*** | .38 | .28 | .51 |
| Female |  |  |  |  |  |
| **A member of family or friend has been confirmed with the disease?** |  |  |  |  |  |
| No | .67 | .29* | 1.96 | 1.12 | 3.43 |
| Yes |  |  |  |  |  |
| **Knew an infected person?** |  |  |  |  |  |
| No | -.90 | .20*** | .41 | .27 | .60 |
| Yes |  |  |  |  |  |
| **Region** |  |  |  |  |  |
| West | .29 | .44 | 1.35 | .57 | 3.17 |
| Center | .52 | .42 | 1.69 | .74 | 3.82 |
| East |  |  |  |  |  |
| **High vs. Low Fear<sup>a</sup>** |  |  |  |  |  |
| Intercept | .379 | .64 |  |  |  |
| **Age** |  |  |  |  |  |
| 18-30 years | -.19 | .34 | .82 | .42 | 1.59 |
| 31-45 years | .22 | .35 | 1.24 | .62 | 2.48 |
| >45 years |  |  |  |  |  |
| **Gender** |  |  |  |  |  |
| Male | -1.51 | .19*** | .22 | .15 | .32 |
| Female |  |  |  |  |  |
| **A member of family or friend has been confirmed with the disease?** |  |  |  |  |  |
| No | .61 | .32 | 1.85 | .99 | 3.46 |
| Yes |  |  |  |  |  |
| **Knew an infected person?** |  |  |  |  |  |
| No | -.98 | .22*** | .38 | .24 | .58 |
| Yes |  |  |  |  |  |
| **Region** |  |  |  |  |  |
| West | -.09 | .53 | .91 | .33 | 2.57 |
| Center | .59 | .50 | 1.81 | .69 | 4.83 |
| East |  |  |  |  |  |

Note: <sup>a</sup>(Reference Category); $R^2 = .09$ (Cox & Snell), $.10$ (Nagelkerke). Model $\chi^2(14) = 108.41, p < 0.001$ . \* $p < 0.05$ ; \*\* $p < 0.01$ ; \*\*\* $p < 0.001$ .

## 4. Discussion

The objective of this study was to explore the relationship between sociodemographic factors and fear of COVID-19 in a Cuban population. Our results suggest that gender and knowing a person positive for COVID-19 significantly increases fear levels in the Cuban population. Several studies have confirmed a greater impact of COVID-19 on the mental health of women compared to men. During the current pandemic, women have shown greater vulnerability to develop higher levels of fear, stress, anxiety, depression, post-traumatic stress symptoms (PTSS), insomnia and adjustment disorder (Wang et al., 2020).

There are also studies conducted with healthcare workers that have shown a greater impact of COVID-19 on the mental health of women compared to men. The results reported greater feelings of uncertainty, fear of contagion, depression, insomnia and a higher incidence of stress reactions among female healthcare workers facing the current COVID-19 outbreak (De Kock et al., 2021).

The consistency of these results emphasizes the need to consider gender in efforts to address the mental health impacts of COVID-19, both in the short term and in the long term (Ahmed & Dumanski, 2020).

Our results also suggest that knowing a person positive for COVID-19 significantly increases fear levels in the Cuban population. Previous studies show that social proximity with people infected with COVID-19 has a significant impact on mental health. For example, having a family member dying of COVID-19 was positively associated with fear of COVID-19 (Bitan et al., 2020).

In another study conducted by Dryhurst et al. (2020), people who have had direct personal experience with the virus perceive more risk compared to those who have not had direct experience; and people who have received information on the virus from family and friends perceive more risk compared to those who have not. A recent study also reported that interpersonal proximity to COVID-19 patients was a predictor variable of mental health (Rossi et al., 2020). The authors reported that in close proximity to infected people was associated with Post-Traumatic Stress Symptom (PTSS), Anxiety and Adjustment Disorder Symptom (ADS). Our results could help authorities of the Ministry of Public Health to design evidence-based intervention strategies, with special emphasis on the most vulnerable groups. For example, there are brief intervention alternatives based on cognitive-behavioral therapy (CBT) that can be used through telepsychology services. It is also important that psychologists in Cuba join the health services where suspected and positive cases of COVID-19 are treated. In this way, mental health can be explored in real time and the necessary interventions carried out with the aim of reducing the negative impact of COVID-19 on mental health. This initiative would not only benefit patients, but also healthcare workers who are on the front lines of the fight against COVID-19. Limitations of the study

This research is not without limitations. First, this is a cross-sectional study so it is difficult to accurately elucidate causal relationships between gender, and interpersonal proximity to COVID-19 patients and fear of COVID-19. Second, our study did not control which participants were in quarantine zones and which were not. Quarantine can amplify negative psychological reactions to COVID-19 (Rossi et al., 2020). On the other hand, although the sample is large, most of the participants (75.1%) correspond to the central area of the country, which implies that the results could probably describe the population of this region of the country in particular.

Additionally, in our sample the presence of mental illnesses was not controlled. Subjects with a mental disorder have been reported to be at higher risk of negative outcomes because of COVID-19 pandemic with respect to the general population (Carmassi et al., 2020).

Conclusions and future directions

In our study, we aimed to explore the relationship between sociodemographic factors and fear of COVID-19 in a Cuban population. We identified that gender and knowing a person positive for COVID-19 significantly increases fear levels in the Cuban population. In future studies, it is important to explore other predictors of the negative impact of COVID-19 on mental health in the Cuban population, such as age, socioeconomic status, exposure to the media (television, digital social networks and print media), the presence of psychiatric comorbidities and non-communicable chronic diseases. Studies should also be conducted to explore the mediating variables of mental health during the pandemic, such as resilience, stress-coping style, among others. These topics of research are very important, especially at the present time, when Cuba is going through the most complex stage of the fight against COVID-19.

## Data Availability

Data is available on http://dx.doi.org/10.17632/3gzdxwcd7y.1

